# Examining the Appeal of Cigarette Filters: An Online Randomised Experiment Among Adults in Great Britain

**DOI:** 10.64898/2026.09.17.26363313

**Authors:** Katherine East, Hazel Cheeseman, Eve Taylor

## Abstract

**Background and aims:** Cigarette filters are present in almost all cigarettes sold globally, despite providing no health protection and contributing substantially to environmental pollution. Filters may also increase cigarette appeal and reinforce misperceptions of reduced harm. This study aimed to examine the impact of cigarette filters on cigarette appeal and perceptions of health and environmental harms among adults in Great Britain.

**Design:** Online between-subject randomised experiment embedded within a nationally representative survey.

**Setting:** Great Britain, May 2026.

**Participants:** Adults aged 18+ years, recruited through the YouGov Social Omnibus survey (n=3961). Subgroup analyses were conducted among people who currently smoked (n=499) and people who smoked daily (n=270).

**Intervention and comparator:** Participants were randomised to view images of cigarettes either without filters (intervention) or with filters (conventional cigarettes; comparator).

**Measurements:** The primary outcome was selecting one of the displayed cigarettes to smoke vs. selecting none. Secondary outcomes were perceptions that the cigarette was extremely harmful to health and extremely harmful to the environment. Logistic regression models were used, adjusting for age, sex and smoking status.

**Results:** There was strong evidence to suggest that participants who viewed cigarettes without filters had markedly lower odds of choosing a cigarette to smoke than those who viewed cigarettes with filters (19.1% vs. 28.1%; adjusted odds ratio [AOR]=0.46 [95% confidence interval [CI]=0.38-0.55], p<.001). Cigarettes without filters also had higher odds of being perceived as extremely harmful to health (65.2% vs. 59.9%; AOR=1.26 [1.10-1.45], p=.001). Evidence was insufficient to conclude a difference between conditions in perceiving cigarettes as extremely harmful to the environment (41.8% vs. 44.3%; AOR=0.89 [0.78-1.01], p=.074). Findings were similar among people who currently smoke, with participants who viewed cigarettes without (vs. with) filters having markedly lower odds of choosing a cigarette (67.4% vs. 94.2%; AOR=0.12 [0.06-0.22], p<.001). These findings were also similar among people who smoke daily (70.7% vs. 94.8%; AOR=0.14, 95% CI=0.06-0.36, p<0.001). However, evidence was insufficient to conclude differences in perceived health harms between conditions among people who smoke currently (29.7% vs. 27.8%; p=.588) or daily (32.2% vs. 28.6%; p=.324).

**Summary:** Removing cigarette filters reduced the appeal of cigarettes among the general population and people who smoke. Removing filters also increased perceptions of health harms in the general population. Given that cigarette filters provide no health protection and contribute to environmental pollution, policies that regulate or prohibit filters may support public health and environmental goals.

## INTRODUCTION

Reducing tobacco smoking remains a UK Government priority.^1,2^ In the UK, over 1 in 10 adults (12.9%) smoke,^3^ with prevalence higher in more disadvantaged groups.^3–5^ Policies are essential for reducing smoking because they can prevent initiation and encourage quitting.^6^ The UK has a strong track record of policies to reduce smoking, and the landmark Tobacco and Vapes Act 2026 provides a unique opportunity to further advance tobacco regulation and improve public health.^1^ Internationally, the World Health Organization’s Framework Convention on Tobacco Control (FCTC) has also recently reported on a number of ‘Forward-Looking Tobacco Control Measures’ to accelerate progress towards reducing smoking.^7^

Cigarette filters have emerged as an increasingly important focus within these discussions. The filter is seen as an integral part of modern cigarettes, with over 90% all cigarettes sold worldwide containing filters.^8^ Expert groups reporting to the FCTC have explored a ban on cigarette filters both on environmental grounds and as a ‘Forward-Looking Tobacco Control Measure’ that could help to reduce smoking. These reports recognise that cigarette filters have no health protection, create significant environmental harms, and remain weakly regulated despite their public health significance.^7^ In the UK, cigarette filters have also been a focus of parliamentary debates during the passage of the Tobacco and Vapes Act.^9–11^ Following an amendment tabled by a cross-party group of parliamentarians to prohibit filters, the Government bought forward its own amendments to create powers to regulate filters in a variety of ways including prohibition. These powers require further regulations in order to be enacted.

The case for regulatory attention is supported by a substantial evidence base. Although the majority of environmental harms of cigarettes comes from tobacco farming and manufacturing,^12^ conventional plastic (cellulose acetate) cigarette filters are the most common form of plastic litter globally and contribute to persistent microplastic pollution.^13^ Yet, people who use them are often unaware that filters contain plastic and synthetic fibers.^14,15^ Decades of research also demonstrate that filters do not reduce the harms associated with smoking; rather, they can facilitate deeper inhalation, expose people who smoke to cellulose acetate fibres and microplastics, and increase cigarette palatability.^8,16–20^ Critically, the widespread availability and use of filters help maintain the longstanding misconception that cigarettes with filters are less harmful than those without.^18,19^

Recent research demonstrates the extent of these misconceptions, finding that over three quarters of adults in Great Britain who smoke incorrectly believe cigarette filters offer protection from the health risks of smoking.^21^ Data from England, Canada, the US, and Australia, further show that 46% of adults who smoke believe that removing filters would make cigarettes more harmful.^22^

Despite growing policy interest, limited research has examined how different cigarette filters influence cigarette appeal.^23^ This is an important evidence gap given the well-established role of product design (e.g., pack design, flavours) in shaping cigarette appeal and use.^24–28^ This study addresses that gap by evaluating the appeal of cigarettes with and without filters, generating timely evidence to inform policy development in the UK and internationally.

This study addresses the following research question: How do cigarette filters impact the appeal of cigarettes? The hypothesis was that more people will choose to smoke cigarettes with filters (i.e., conventional cigarettes) than cigarettes without filters. The impact of cigarette filters on harm and environmental perceptions were also explored.

## METHODS

### Public involvement and pre-registration

Two adults with lived or living experience with cigarette smoking informed the survey and images used. This study was also pre-registered (<u>osf.io/pb9wf</u>)^29^ and the data and code are available online at https://osf.io/8hk2j/files/osfstorage.

### Design

This study used an online random experiment embedded into a national survey. The experiment had two conditions: (1) cigarettes with filters (i.e., conventional cigarettes); (2) cigarettes without filters. See Figure 1.

**Figure 1.**
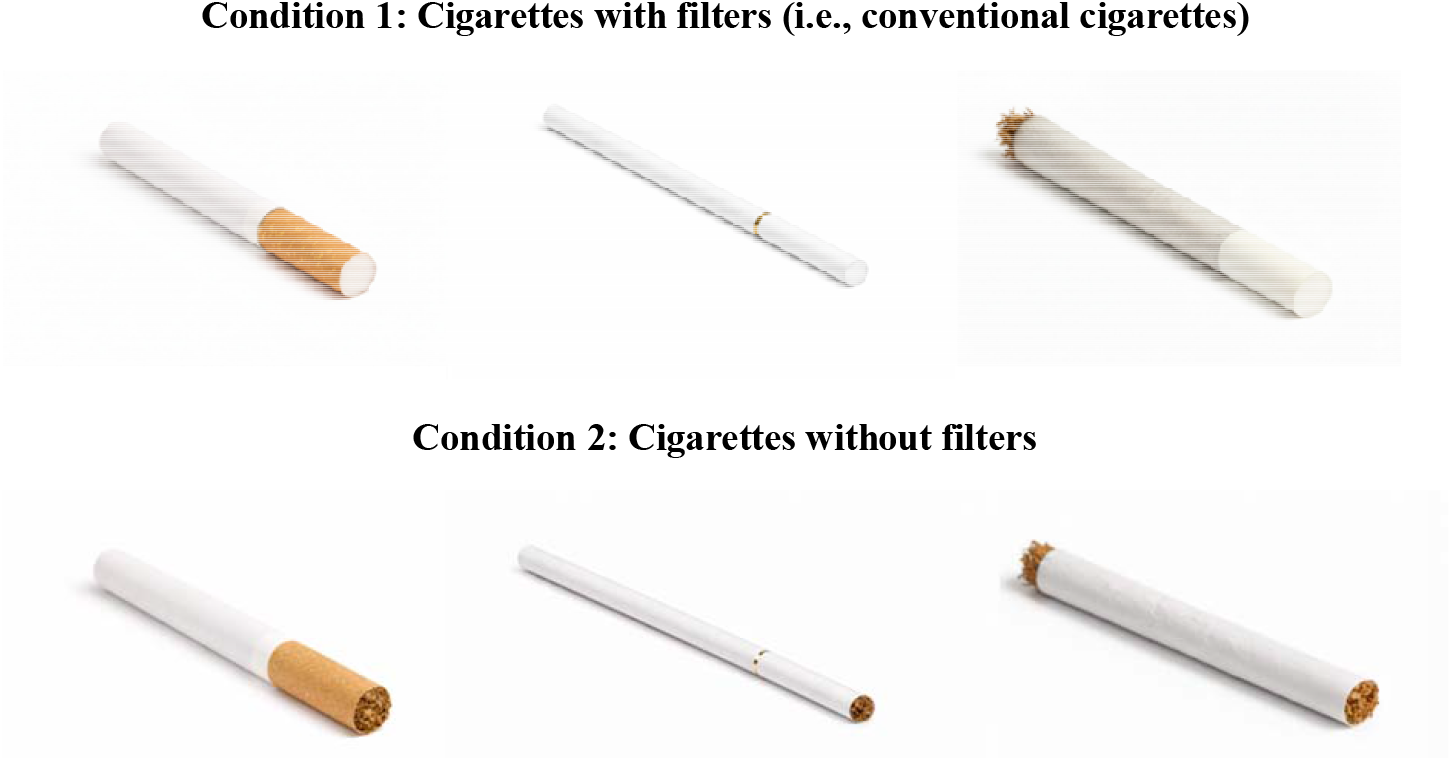
Conditions used in the experiment.

### Recruitment and sample

Data were collected by YouGov via their Social Omnibus between 14^th^ and 25^th^ May 2026. YouGov’s Social Omnibus recruits ~2,000 adults aged 18+ across Great Britain per survey, with samples designed to be nationally representative. This study ran over two survey days, sampling a total of n=4,019 adults who consented to take part. Respondents were excluded listwise from all analyses where they responded “prefer not to say” on key variables (n=58), resulting in an analytical sample of n=3,961.

### Measures

The survey measures including routing are available online (osf.io/pb9wf).^29^

#### Primary outcome (choosing to smoke any of the cigarettes displayed)

Respondents were shown a set of 3 cigarettes per condition and asked, “If these were the only cigarettes available to you, which one would you choose to smoke, if any?” Response options included selecting one of the cigarettes displayed, “I would not smoke any of these cigarettes”, “Don’t know”, and “Prefer not to say”. Responses were categorised as selected a cigarette vs. “I would not smoke any of these cigarettes”. “Don’t know” (n=55) and “Prefer not to say” (n=9) responses were excluded. This approach is consistent with our prior work.^30–32^

#### Secondary outcomes (harm perceptions, environmental perceptions)

Within each condition, participants were shown one image at random and asked (1) “How harmful do you think it is to smoke this cigarette?” and (2) “How harmful do you think this cigarette is to the environment?”. For both, response options were on a sliding scale from 0 (not at all harmful) to 10 (extremely harmful), with additional options “Don’t know” and “Prefer not to say”. Responses were coded as “extremely harmful” vs. otherwise (including “Don’t know”). Respondents who select “Prefer not to say” were excluded (n=9). As a sensitivity analysis, mean scores were compared, excluding “Don’t know” and “Prefer not to say” responses.

#### Independent variable (condition)

Cigarettes with filters (i.e., conventional cigarettes) vs. cigarettes without filters.

#### Covariates

For the main analyses, covariates were sex (male, female), age group (18-29, 30-39, 40-49, 50-59, 60+), and smoking status (never, former, current; with “Don’t know” and “Prefer not to say” responses excluded from analyses [n=47]). For subgroup analyses among people who currently smoke, covariates were sex, age group, and type of cigarette smoked (only/mainly factory-made/pack, otherwise; with “Don’t know” and “Prefer not to say” responses excluded from analyses [n=10]). For subgroup analyses among people who smoke daily, covariates were sex, age group, and heaviness of smoking (low, medium/high; with “Don’t know” and “Prefer not to say” responses excluded from analyses [n=8]).

### Analyses

Between-subject analyses were used. For each outcome, unadjusted and adjusted (for all covariates) binary logistic regressions were run with condition as the independent variable. For the sensitivity analyses for the harm perceptions and environmental perceptions (i.e., secondary) outcomes coded as continuous, linear and ordinal regressions were run with condition as the independent variable, adjusting for covariates. Analyses were first run among the whole sample, followed by subgroup analyses among people who currently smoke and people who smoke daily, adjusting for covariates. Effects were reported with 95% confidence intervals and p values.

The following adjustments were made from the pre-registered analyses. First, we planned to treat “don’t know” responses to the primary outcome (choosing to smoke any of the cigarettes displayed) as a unique level and run multinomial regressions; however, due to small numbers of participants who selected “don’t know” (n=55), we excluded this response option and ran binary logistic regressions. Second, for sensitivity analyses, we planned to run linear regressions only; however, assumptions checks indicated that residuals may not normally distributed; therefore ordinal logistic regression analyses were also run to produce more robust findings. Third, subgroup analyses were planned among people who currently smoke; however, heaviness of smoking was only asked among people who smoke daily, so subgroup analyses were also run among this group to control for heaviness of smoking.

## RESULTS

Participant characteristics are displayed in Table 1 and smoking characteristics in Table S1. Randomisation checks indicated a possible difference in age distribution between participants in the two conditions (p=.049), but no difference in gender or smoking status (p>.05; Table 1).

**Table 1:** Participant characteristics by experimental condition (n=3961).

| | All participants | Experimental condition | | $\chi^2$ P |
| --- | --- | --- | --- | --- |
|  |  | With filter | No filter |  |
| Age |  |  |  |  |
| 18-24 | 420(10.6%) | 232 (55.2%) | 188 (44.8%) | .049 |
| 25-34 | 717(18.1%) | 341 (47.6%) | 376 (52.4%) |  |
| 35-44 | 683(17.2%) | 339 (49.6%) | 344 (50.4%) |  |
| 45-54 | 743(18.8%) | 350 (47.1%) | 393 (52.9%) |  |
| 54-64 | 574(14.5%) | 264 (46.0%) | 310 (54.0%) |  |
| 65+ | 824(20.8%) | 416 (50.5%) | 408 (49.5%) |  |
| Gender |  |  |  |  |
| Male | 1907(48.1%) | 951 (49.9%) | 956 (50.1%) | .308 |
| Female | 2054(51.9%) | 991 (48.2%) | 1,063 (51.8%) |  |
| Smoking status |  |  |  |  |
| Never | 2091(52.8%) | 1,025 (49.0%) | 1,066 (51.0%) | .928 |
| Former | 1371(34.6%) | 676 (49.3%) | 695 (50.7%) |  |
| Current | 499(12.6%) | 241 (48.3%) | 258 (51.7%) |  |

### Choosing a cigarette

In unadjusted and adjusted analyses, participants who viewed cigarettes without filters had lower odds of choosing a cigarette to smoke compared to those who viewed cigarettes with filters (19.1% vs. 28.1%; AOR=0.46 [95% CI=0.38-0.55], p<.001; Table 2). Choosing a cigarette was also greater among those who currently or formerly smoked compared to those who never smoked, those aged 18-24 compared to those aged over 45, and among males compared to females (Table 2).

**Table 2.** Associations between choosing a cigarette, perceptions of harm to health, the environment, and filter condition.

|  | Choosing (vs. not choosing) a cigarette<br>(n=3906) <sup>1</sup> |  |  | Perceiving cigarette as extremely harmful to<br>health (vs. otherwise, n=3961) |  |  | Perceiving cigarette as extremely harmful to<br>the environment (vs. otherwise, n=3961) |  |  |
| --- | --- | --- | --- | --- | --- | --- | --- | --- | --- |
|  | n/N (%)1 | AOR (95% CI) | p | n/N (%) | AOR (95% CI) | p | n/N (%) | AOR (95% CI) | p |
| **Experimental condition** |  |  |  |  |  |  |  |  |  |
| With filter | 539/1,918 (28.1%) | 1 | Ref | 1,164/1,942 (59.9%) | 1 | Ref | 860/1,942 (44.3%) | 1 | Ref |
| No filter | 380/1,988 (19.1%) | 0.46 (0.38-0.55) | <.001 | 1,316/2,019 (65.2%) | 1.26 (1.10-1.45) | .001 | 844/2,019 (41.8%) | 0.89 (0.78-1.01) | .074 |
| **Smoking status** |  |  |  |  |  |  |  |  |  |
| Never | 130/2,075 (6.3%) | 1 | Ref | 1,497/2,091 (71.6%) | 1 | Ref | 1,098/2,091 (52.5%) | 1 | Ref |
| Former | 410/1,353 (30.3%) | 8.49 (6.77-10.65) | <.001 | 838/1,371 (61.1%) | 0.53 (0.46-0.62) | <.001 | 524/1,371 (38.2%) | 0.51 (0.44-0.59) | <.001 |
| Current | 379/478 (79.3%) | 68.44 (50.78-92.24) | <.001 | 145/499 (29.1%) | 0.16 (0.13-0.20) | <.001 | 82/499 (16.4%) | 0.18 (0.14-0.23) | <.001 |
| **Age** |  |  |  |  |  |  |  |  |  |
| 18-24 | 109/414 (26.3%) | 1 | Ref | 207/420 (49.3%) | 1 | Ref | 143/420 (34.0%) | 1 | Ref |
| 25-34 | 199/709 (28.1%) | 1.12 (0.78-1.61) | .546 | 407/717 (56.8%) | 1.40 (1.08-1.80) | .011 | 287/717 (40.0%) | 1.37 (1.05-1.77) | .019 |
| 35-44 | 183/673 (27.2%) | 0.82 (0.57-1.18) | .282 | 424/683 (62.1%) | 1.93 (1.49-2.51) | <.001 | 309/683 (45.2%) | 1.85 (1.42-2.41) | <.001 |
| 45-54 | 175/731 (23.9%) | 0.66 (0.46-0.95) | .026 | 473/743 (63.7%) | 2.03 (1.57-2.63) | <.001 | 316/743 (42.5%) | 1.63 (1.26-2.12) | <.001 |
| 54-64 | 125/564 (22.2%) | 0.59 (0.40-0.87) | .007 | 394/574 (68.6%) | 2.57 (1.94-3.39) | <.001 | 260/574 (45.3%) | 1.86 (1.42-2.45) | <.001 |
| 65+ | 128/815 (15.7%) | 0.32 (0.22-0.47) | <.001 | 575/824 (69.8%) | 2.67 (2.05-3.47) | <.001 | 389/824 (47.2%) | 2.00 (1.55-2.60) | <.001 |
| **Gender** |  |  |  |  |  |  |  |  |  |
| Male | 511/1,883 (27.1%) | 1 | Ref | 1,090/1,907 (57.2%) | 1 | Ref | 713/1,907 (37.4%) | 1 | Ref |
| Female | 408/2,023 (20.2%) | 0.70 (0.58-0.84) | <.001 | 1,390/2,054 (67.7%) | 1.52 (1.33-1.75) | <.001 | 991/2,054 (48.2%) | 1.52 (1.33-1.73) | <.001 |
| **Unadjusted model** |  |  |  |  |  |  |  |  |  |
| **Experimental condition** |  |  |  |  |  |  |  |  |  |
| With filter | 539/1,918 (28.1%) | 1 | Ref | 1,164/1,942 (59.9%) | 1 | Ref | 860/1,942 (44.3%) | 1 | Ref |
| No filter | 380/1,988 (19.1%) | 0.60 (0.52-0.70) | <.001 | 1,316/2,019 (65.2%) | 1.25 (1.10-1.42) | .001 | 844/2,019 (41.8%) | 0.90 (0.80-1.02) | .115 |

1 Excludes people who responded “Don’t know” or “Prefer not to say” to choosing a cigarette (n=55).
AOR=adjusted odds ratio; 95% CI=95% confidence interval; ref=reference category.
**Bold** denotes p<.05.

Findings were consistent in the subgroup analyses (Table 3), such that those who currently smoked and viewed cigarettes without filters had lower odds of choosing a cigarette than those who currently smoked and viewed cigarettes with filters (67.4% vs. 92.3%; AOR=0.12 [0.06-0.22], p<.001), after adjusting for age, gender, and type of cigarette usually smoked. Findings were similar among those who smoked daily, after adjusting for age, gender, and heaviness of smoking (70.7% vs. 94.8%; AOR=0.14 [0.06-0.36], p<.001).

**Table 3.** Subgroup analyses: associations between choosing a cigarette, perceptions of harm to health, the environment and filter condition among people who smoke.

|  | Choosing (vs. not choosing) a cigarette |  |  | Perceiving cigarette as extremely harmful to health (vs. otherwise) |  |  | Perceiving cigarette as extremely harmful to the environment (vs. otherwise) |  |  |
| --- | --- | --- | --- | --- | --- | --- | --- | --- | --- |
|  | n/N (%) | AOR (95 % CI) | p | n/N (%) | AOR (95 % CI) | p | n/N (%) | AOR (95 % CI) | p |
| **People who currently smoke, including adjustment for cigarette type <sup>1,2</sup>** |  |  |  |  |  |  |  |  |  |
| **Experimental condition** |  |  |  |  |  |  |  |  |  |
| With filter | 213/226(94.2%) | 1 | Ref | 65/234(27.8%) | 1 | Ref | 41/234(17.5%) | 1 | Ref |
| No filter | 163/242(67.4%) | 0.12(0.06-0.22) | **<.001** | 76/255(29.8%) | 1.12(0.75-1.67) | .588 | 38/255(14.9%) | 0.64(0.39-1.05) | .079 |
| **Type of cigarette usually smoked** |  |  |  |  |  |  |  |  |  |
| Only/mainly factory-made | 222/268 (82.8%) | 1 | Ref | 73/281(26.0%) | 1 | Ref | 53/281(18.9%) | 1 | Ref |
| Other | 154/200(77.0%) | 0.53(0.32-0.89) | **.016** | 68/208(32.7%) | 1.34(0.90-2.01) | .154 | 26/208(12.5%) | 0.61(0.36-1.02) | .061 |
| **People who smoke daily, including adjustment for heaviness of smoking <sup>3,4</sup>** |  |  |  |  |  |  |  |  |  |
| **Experimental condition** |  |  |  |  |  |  |  |  |  |
| With filter | 110/116(94.8%) | 1 | Ref | 34/119(28.6%) | 1 | Ref | 16/119(13.4%) | 1 | Ref |
| No filter | 94/133 (70.7%) | 0.14(0.06-0.36) | **<.001** | 46/143(32.2%) | 0.76(0.44-1.32) | .324 | 17/143(11.9%) | 0.42(0.19-0.91) | **.029** |
| **Heaviness of smoking index** |  |  |  |  |  |  |  |  |  |
| Low | 113/136(83.1%) | 1 | Ref | 45/147(30.6%) | 1 | Ref | 19/147(12.9%) | 1 | Ref |
| Moderate/high | 91/113(80.5%) | 1.13(0.56-2.30) | .729 | 35/115(30.4%) | 1.00(0.58-1.72) | .986 | 14/115(12.2%) | 1.05(0.49-2.26) | .897 |

<sup>1</sup> Adjusted for age, gender, and cigarette type.
<sup>2</sup> Excludes people who responded “Don’t know” or “Prefer not to say” to type of cigarette smoked (n=10) and, for the model with choosing a cigarette as the outcome, also excludes people who responded “Don’t know” or “Prefer not to say” to choosing a cigarette (n=21).
<sup>3</sup> Adjusted for age, gender, and heaviness of smoking.
<sup>4</sup> Excludes people who responded “Don’t know” or “Prefer not to say” to heaviness of smoking (n=8) and, for the model with choosing a cigarette as the outcome, also excludes people who responded “Don’t know” or “Prefer not to say” to choosing a cigarette (n=14).
AOR=adjusted odds ratio; 95% CI=95% confidence interval; ref=reference category.
**bold** denotes p<.05.

### Perceptions of harm to health

In both unadjusted and adjusted analyses, participants who viewed cigarettes without filters had greater odds of perceiving the cigarette displayed as extremely harmful to health than those who viewed cigarettes with filters (65.2% vs. 59.9%; AOR=1.26[1.10-1.45], p=.001; Table 2). Odds of perceiving the cigarette displayed as extremely harmful to health were lower among those who currently or formerly smoked compared to never smoked, among those aged 18-24 compared to all older age groups, and among males compared to females (Table 2).

Unlike the main analyses, subgroup analyses found insufficient evidence of a difference between conditions in perceiving that the cigarette displayed is extremely harmful to health, among those who smoke currently (adjusting for age, gender, and type of cigarette smoked) or daily (adjusting for age, gender, and heaviness of smoking; Table 3).

Findings for the sensitivity analyses using ordinal and linear regressions were consistent with the main analyses, such that participants perceived greater health harms when viewing cigarettes without a filter compared to cigarettes with a filter (original: AOR=1.18[1.03-1.35], p=.015; linear: adjusted beta=0.10[0.01-0.19], p=.037; Supplementary Table S2; Figure S1).

### Perceptions of harm to the environment

In both adjusted and unadjusted analyses, there was insufficient evidence of a difference in perceiving the cigarette displayed as extremely harmful to the environment between those who viewed cigarettes without (vs. with) filters (41.3% vs. 44.8%; AOR=0.89[0.78-1.01], p=.074; Table 2). Odds of perceiving the cigarette displayed as extremely harmful to the environment were lower among those who currently or formerly smoked compared to those who never smoked, among those aged 18-24 compared to all older age groups, and among males compared to females (Table 2).

Findings were consistent in the subgroup analyses among people who currently smoke, adjusting for age, gender, and cigarette type, such that there was insufficient evidence of a difference between conditions in perceiving the cigarette displayed as extremely harmful to the environment (Table 3). However, among people who smoke daily in analyses adjusted for age, gender, and cigarette type, those in the condition without (vs. with) filters had lower odds of perceiving the cigarette displayed as harmful to the environment (11.9% vs. 13.4%; AOR=0.42[0.19-0.91], p=.029; Table 3).

Sensitivity analyses using ordinal and linear regressions indicated that participants perceived lower environmental harms when viewing a cigarette in the condition without (vs. with) filters (ordinal: AOR=0.75 [0.67-0.85], p<.001; linear: adjusted beta=-0.46[-0.61--0.30, p<.001) (Supplementary Table S2; Figure S1).

## DISCUSSION

As hypothesised, more adults in Great Britain chose to smoke cigarettes with filters (i.e., conventional cigarettes) than cigarettes without filters, including adults who smoke. This suggests that removing filters reduces the appeal of cigarettes. Adults also perceived that cigarettes were more harmful to health if they did not contain filters; however, this was not observed among people who smoke. The main analyses found insufficient evidence for differences in harm perceptions to the environment; however, sensitivity analyses indicated that cigarettes without filters were perceived as less environmentally harmful than those with filters.

This is the first independent study, to our knowledge, to assess the impact of cigarette filters on the appeal of cigarettes, as well as considering health and environmental perceptions. The findings extend prior research demonstrating widespread misperceptions that filters reduce the health harms from smoking.^18,19,21,22^ Our findings further suggest that filters influence not only perceptions but also behavioural intentions, with participants substantially less likely to choose a cigarette when filters were removed. Together, these findings support the argument that filters serve as a product design feature that increase appeal while conveying misleading cues about risk.^17,21,23^

The effect among people who smoke was particularly prominent, with 94% choosing a cigarette with a filter compared to 67% choosing a cigarette without a filter, representing an approximately 80% reduction in the odds of choosing a cigarette. This is consistent with research finding that people who smoked cigarettes with (vs. without) filters rated them as better tasting, more satisfying, and enjoyable.^19^ It is also consistent with wider research on tobacco product design, such as standardised packaging which has demonstrated reductions in cigarette appeal.^33,34^ Future research should explore whether removal of cigarette filters predicts actual behaviour, such as reductions in uptake of smoking and increases in quitting.

While cigarettes without filters were viewed as more harmful to health among the general population, consistent with prior research,^18,19,21,22^ this difference was not observed among adults who smoke. This may be due to the much lower overall perceptions of harm from smoking among adults who smoke compared to the general population, as was seen in this study, or more entrenched beliefs about smoking harms that are less influenced by product design. Nevertheless, adults who smoked remained less likely to choose cigarettes without filters, suggesting that filters may influence behavioural intentions through mechanisms beyond harm perceptions.

Removing filters also had little effect on perceptions of environmental harm in the main analyses, despite cigarette filters being the most common form of plastic litter globally.^13^ However, in subgroup analyses among people who smoke daily and sensitivity analyses using a more granular analytic approach, cigarettes without filters were perceived as less environmentally harmful than those with filters. This may indicate some awareness of the environmental harms associated with filters, consistent with findings among US youth and young adults, in which 89% agreed that filters are harmful to the environment.^15^ One concern is that reduced environmental harm perceptions could lead to increased cigarette littering. Further research is therefore required to assess the impact of removing cigarette filters on littering behaviours and overall environmental impact.

The dual findings that removing cigarette filters increased perceptions of health harms but had little effect on environmental perceptions among the general population suggest that any regulatory action on filters may need to be accompanied by public communication explaining the public health and environmental rationale.^23^ Both health and environmental messaging could help counter the perception that filters are a benign or beneficial component of cigarettes. This could be in the form of public education campaigns, packaging labels, pack inserts, or messages on the cigarette sticks/filters themselves.^23^ Further research is required to identify which messages would work, and in what format.

The findings have important policy implications. Overall, they suggest that removing cigarette filters could reduce cigarette appeal while simultaneously addressing misleading perceptions of reduced harm. While many tobacco product regulations focus on reducing uptake, a filter ban shows promise for reducing appeal among adults who smoke. A filter ban has yet to be implemented in any country, although Santa Cruz (a US city) finalised a bill banning cigarette and cigar filters in October 2024, planned for 2027; this is set to be the first globally. Filter bans have also been recommended by government bodies in Belgium, considered by the Dutch government,^35,36^ and been the focus of parliamentary debates in the UK.^9–11^ The UK Tobacco and Vapes Act provides a unique opportunity for further restrictions on filters and consideration of a filter ban, as it provides Government with wide-ranging regulatory powers.^21^

This study has limitations. First, participants responded to images of cigarettes rather than making real decisions about purchasing or use of cigarettes, and behavioural intentions may not translate into real-world behaviour. Second, the experiment measured immediate responses and findings may not translate to long-term behaviour changes or adaptation to the removal of filters. Third, the study was conducted among adults in Great Britain, so findings may not generalise to countries with different tobacco markets and policies and tell us nothing about how filters impact appeal to young people. However, strengths of this study include that it was pre-registered, used a randomised design to strengthen causal inference, used a large, nationally recruited sample, and considered cigarette filter appeal among the general population as well as adults who smoke.

In conclusion, removing cigarette filters substantially reduced the appeal of cigarettes, particularly among adults who smoke, while increasing perceptions that smoking is harmful. Given that filters offer no health protection and create considerable environmental harms, these findings provide timely evidence to inform current UK and international discussions on cigarette filter regulation. Removing cigarette filters may represent a novel policy option capable of reducing cigarette appeal while addressing both public health and environmental goals.

## Data Availability Statement

The data and code behind this work are available online at https://osf.io/8hk2j/files/osfstorage.

