## Supplementary File for "Examining the Appeal of Cigarette Filters: An Online Randomised Experiment Among Adults in Great Britain"

| **Table S1: Smoking characteristics.** | |  |
| --- | --- | --- |
|  | **People who currently smoke (n=499) n(%)** | **People who smoke daily (n=270) n(%)** |
| **Frequency of smoking** |  |  |
| Daily | 270(54.1) | - |
| Non-daily | 229(45.9) | - |
| **Cigarette type smoked^1^** | |  |
| Only/mainly factory-made | 281(57.5%) |  |
| Other | 208(42.5%) |  |
| **Heaviness of smoking index*^2^*** | |  |
| Low heaviness of smoking | - | 147(56.1%) |
| Moderate/high heaviness of smoking | - | 115(43.9%) |
| ^1^ Excludes people who responded “Don’t know” or “Prefer not to say” to cigarette type smoked (n=10).  ^2^ Not assessed among people who do not smoke daily. Excludes people who responded “Don’t know” or “Prefer not to say” to heaviness of smoking (n=8). | | |

| **Table S2: Sensitivity analysis assessing ordinal and linear associations between perceptions of harm to health and the environment, and filter condition.** | | | | |
| --- | --- | --- | --- | --- |
|  | **Health harm^3^** | | **Environmental harm^4^** | |
| **Ordinal regression^1^** | **AOR (95% CI)** | **p** | **AOR (95% CI)** | **p** |
| **Experimental condition** | |  |  |  |
| With filter | 1 | Ref | 1 | Ref |
| No filter | 1.18 (1.03-1.35) | **.015** | 0.75 (0.67-0.85) | **<.001** |
| **Linear regression^2^** | **Adjusted Beta (95% CI)** | **p** | **Adjusted Beta (95% CI)** | **p** |
| **Experimental condition** | |  |  |  |
| With filter | 0 | Ref | 0 | Ref |
| No filter | 0.10 (0.01-0.19) | **.037** | -0.46 (-0.61--0.30) | **<.001** |
| ^1^ Adjusted for age, gender and smoking status. Ordinal logistic regression ORs represent the odds of reporting a response in a higher outcome category; ORs >1 indicate higher perceived harm and ORs <1 indicate lower perceived harm.  ^2^ Adjusted for age, gender and smoking status. Linear regression coefficients represent the adjusted mean difference in outcome score relative to the reference category; positive coefficients indicate higher perceived harm and negative coefficients indicate lower perceived harm.  ^3^ Excludes people who responded “Don’t know” or “Prefer not to say” to health harm (n=131).  ^4^ Excludes people who responded “Don’t know” or “Prefer not to say” to environmental harm (n=375).  AOR=adjusted odds ratio; 95% CI=95% confidence interval; ref=reference category.  **Bold** denotes p<.05. | | | | |

Figure S1: Perceived harm of cigarettes to health and the environment by filter condition (N=3961)

%

|  | 0 (not at all harmful) | 1 | 2 | 3 | 4 | 5 | 6 | 7 | 8 | 9 | 10 (extremely harmful) | Don’t know |
| --- | --- | --- | --- | --- | --- | --- | --- | --- | --- | --- | --- | --- |
| Health Harms With filter | 0.26 | 0.15 | 0.36 | 0.36 | 0.77 | 2.83 | 2.88 | 7 | 12 | 9.37 | 59.94 | 4.07 |
| Health Harms Without filter | 0.25 | 0.15 | 0.15 | 0.5 | 0.54 | 2.08 | 2.82 | 7.13 | 11.24 | 7.38 | 65.18 | 2.58 |
| Enviromental harms With filter | 0.82 | 0.77 | 1.34 | 1.75 | 1.85 | 5.61 | 5.15 | 10.09 | 11.12 | 7.72 | 44.28 | 9.47 |
| Enviromental harms Without filter | 2.38 | 1.88 | 1.98 | 2.77 | 2.13 | 6.24 | 6.39 | 9.01 | 10.5 | 5.45 | 41.8 | 9.46 |
